# Electrocardiogram Analysis of Post-Stroke Elderly People Using One-dimensional Convolutional Neural Network Model with Gradient-weighted Class Activation Mapping

**DOI:** 10.1101/2021.09.29.21264316

**Authors:** Eric S. Ho, Zhaoyi Ding

## Abstract

**Background and purposes:** Stroke is the second leading cause of death globally after ischemic heart disease, also a risk factor of cardioembolic stroke. Thus, we postulate that heartbeats encapsulate vital signals related to stroke. With the rapid advancement of deep neural networks (DNNs), it emerges as a powerful tool to decipher intriguing heartbeat patterns associated with post-stroke patients. In this study, we propose the use of a one-dimensional convolutional network (1D-CNN) architecture to build a binary classifier that distinguishes electrocardiogram s (ECGs) between the post-stroke and the stroke-free.

**Methods:** We have built two 1D-CNNs that were used to identify distinct patterns from an openly accessible ECG dataset collected from elderly post-stroke patients. In addition to prediction accuracy, which is the primary focus of existing ECG deep neural network methods, we have utilized Gradient-weighted Class Activation Mapping (GRAD-CAM) to ease model interpretation by uncovering ECG patterns captured by our model.

**Results:** Our stroke model has achieved ∼90% accuracy and 0.95 area under the Receiver Operating Characteristic curve. Findings suggest that the core PQRST complex alone is important but not sufficient to differentiate the post-stroke and the stroke-free.

**Conclusions:** We have developed an accurate stroke model using the latest DNN method. Importantly, our work has illustrated an approach to enhance model interpretation, overcoming the black-box issue facing DNN, fostering higher user confidence and adoption of DNN in medicine.

## Introduction

Stroke is the second and the fifth leading cause of death globally and in the United States, respectively[1, 2]. It is also a major cause of long-term disability for adults (3.1% of adults)[3]. WHO global statistics reported that stroke has elevated from the fifth-highest to the third-highest disability-adjusted life years (DALY) globally, reaching 139 million in 2019 from 126 million in 2000[4]. Besides, stroke widens health disparity between high-income and low-income countries where “70% of strokes and 87% of both stroke-related deaths and disability-adjusted life-years occur in low-income and middle-income countries” [5]. Cardioembolism is the leading stroke subtype in whites (28%) according to a meta-analysis of stroke subtypes between 1993 and 2015[6]. It is also the second largest ischemic stroke subtype in South Korea[7]. Alarmingly, cardioembolism is on a rising trend in both whites and Koreans[5-7]. It is also linked to ischemic heart disease among Chinese in Taiwan[8]. Thus, giving the pan-population impact, studying heartbeat patterns offers a promising avenue to understand the impact of cardiac activities after stroke.

Deep neural network (DNN) is a transformative artificial intelligence method that brings leap-frog development to medicine and numerous other areas. A DNN architecture is called convolutional neural network (CNN), which is particularly designed for processing time series data such as electrocardiograms (ECGs). Previous studies had used CNNs to detect ECG characteristic waveforms[9], arrhythmia[10, 11], atrial fibrillation[12, 13], hypertrophic cardiomyopathy[14], and myocardial infarction[15].

Here we propose the use of a one-dimensional convolutional network (1D-CNN) architecture to build a binary classifier that distinguishes ECGs between the post-stroke and the stroke-free. While DNNs have gained an unprecedented reputation in solving complex problems, outperformed existing machine learning methods, features or patterns leveraged by the DNN model are often largely unknown, hindering interpretation. It particularly worries clinicians in making therapeutic decisions. Thus, in the present study, we have taken extra steps to unravel ECG patterns our model has harnessed in discerning the two classes of subjects, in addition to prediction accuracy, such that clinicians, as well as patients, are confident of its benefit.

## Materials and Methods

### ECG Data

This study was based on an openly accessible ECG dataset from the study, “Cerebral Vasoregulation in Elderly with Stroke”[16], acquired from PhysioNet[17, 18]. The study aimed to investigate the effects of ischemic stroke on cerebral vasoregulation. Here we provide a brief synopsis of the study protocol. Readers should consult the full study protocol document (day1-day2-protocol.docx) for details from the study’s website[19]. This study recruited 60 post-stroke and 60 stroke-free subjects from the greater Boston area, but only 43 stroke and 48 stroke-free subjects had completed the study. Subjects were 60 to 80 years of age, with a nearly even gender distribution (49 females vs. 42 males). The stroke group included subjects with the first hemispheric ischemic stroke with documented neurological deficits persisting longer than 24 hours, and they were at least six months after stroke. Stroke etiology includes large vessel intracranial atherosclerosis (18 subjects), cardioembolism (5 subjects), atherothrombosis (5 subjects), and undetermined/unknown (16 subjects). Subjects in the control group had been examined with no clinical history of stroke and focal deficit. Subjects were further stratified as “hypertensive” (BP ≥ 140/90 mmHg) and “normotensive” (BP < 140/100) with no blood pressure-controlling medications. The protocol lasted for two days with an overnight stay in General Clinical Research Center (GCRC). Each day, a subject was scheduled to participate in various activities according to a timetable, such as active standing, walking, eating, and sleeping.

### ECG Data Preprocessing

The ECG dataset obtained from PhysioNet was captured by ME6000 device (MegaElectronics) at 1000 Hz. ECGs were measured using a 3-lead electrocardiogram, digitalized, and recorded in WFDB format[20]. To facilitate processing, we utilized the wfdb package (3.3.0)[21] to convert digitalized signals recorded in channels 1 (V5/6-L clavicle) and 2 (V1/2-L clavicle) into plain text files, namely .p_signal files. As the ECG of each subject was recorded in two to three separate files, 402 .p_signal files were created from 91 subjects.

In the original study, the ECG of each subject covered approximately 24 hours, equivalent to 86.4 million milliseconds (ms) per subject. We decided to sample a small fraction of the ECG data to form the learning dataset for two reasons: i) the sheer volume is too large (91 subjects times 86.4 million ms) for our computer hardware, and ii) the original ECG data was not annotated by activity, we were unable to identify the activity (e.g. walking, sleeping, etc.) associated with each time period. Our solution was to randomly sample one hundred non-overlapping segments, each 6000-ms long, per .p_signal file but skipping the first and the last half an hour. Some .p_signal files were found to cover only a short period; as a result, only 34,200 segments were selected. This formed the initial learning dataset for building our stroke model.

It is noteworthy that different segment lengths were considered, such as 2,000 ms, 4,000 ms, and 10,000 ms. From the perspective of model accuracy and interpretability, 6,000 ms was the length chosen for this project. Refer to the Results section for more details about the effect of segment length on model performance.

### ECG Normalization

To ensure that network weights converge during training, signal values from .p_signal files were normalized and stored in separate files, namely .normalized files. The normalization was done using MinMaxScaler method as described below:

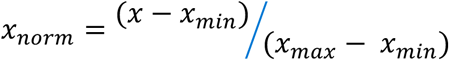

where *x*_*min*_ and *x*_*max*_ are the minimum and maximum values in the segment, respectively. As a result, raw ECG electrical signals (x’s) were rescaled to the range between 0 and 1.

### Derived ECG Features

Four features were derived from the normalized segments: smoothed signal, high-pass signal, the first derivative of the smoothed signal, and the second derivative of the smoothed signals (Figure 1A).

**Figure 1.**
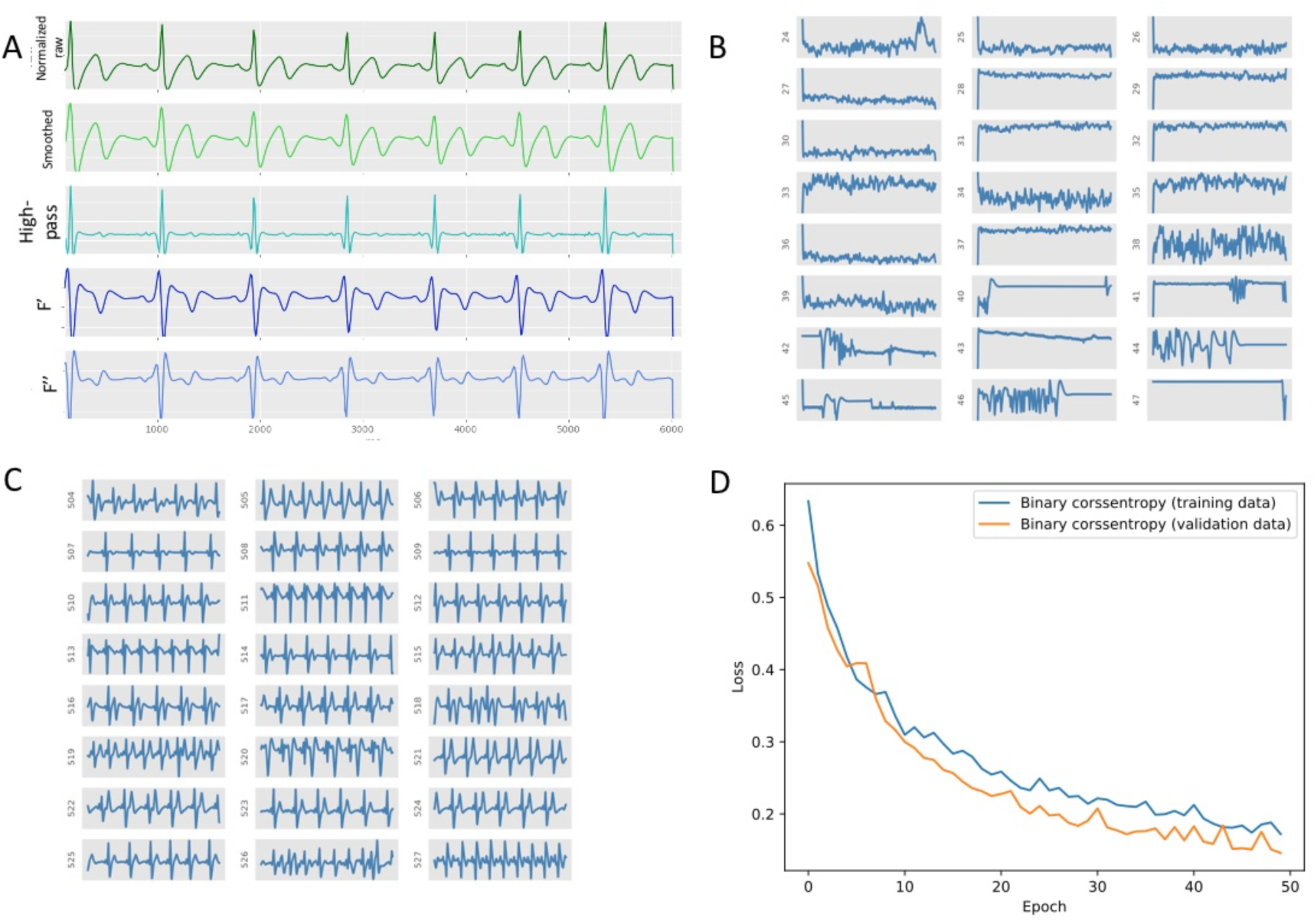
A. Pre-processing of raw digitized ECG data and four features, including the smoothed data, high-pass data, the first and second derivatives of the smoothed data. B-C. The left and right panels at the top display 24 samples of low- and high-quality ECG segments, respectively. D. The plot shows the training and validation losses versus epoch. As you can see in the plot decreases steadily, indicating the absence of overfitting.

To remove small random fluctuations from the normalized raw signals, we used a low-pass filter to drop them out by the scipy’s butter package (v1.6.3)[22] that implemented the Butterworth filter method. The cutoff frequency and polynomial order were 11 Hz and two (quadratic), respectively. As high-frequency signals may also be informative, we isolated high-frequency signals by subtracting the smoothed signals from the normalized signals. The rate at which the signals change may complement the smoothed and high-frequency features. Therefore, the first and second derivatives of the smoothed signal were computed. The first derivative was estimated by an imaginary slope triangle, with a base length of ten units, sliding along the time axis point by point. The slope equaled the y-value of the tenth data point subtracted from the value of the first data point and divided by ten. The second derivative was estimated similarly, except that the input data was the first derivative of the signals.

### Quality Checking Model

We set to remove low-quality signals from the learning dataset before using them to train the stroke model. A handful of low-quality signals are depicted in Figure 1B. To clean the data, we built a quality checking (QC) one-dimensional CNN model to distinguish high-quality signals from low-quality segments, namely a binary classifier, such that only high-quality segments were retained for developing the stroke model.

A semi-automatic approach was taken to compile the learning dataset as below:

- Four non-overlapping 6000-ms segments were randomly selected from each subject, creating 1,608 segments.
- A custom Python script was developed to plot those segments, as shown in Figure 1B-C.
- Segments were manually labeled as low- or high-quality by visual inspection of plots generated from the previous step.

In the end, the learning data comprised of 476 and 443 low- and high-quality segments, respectively. An 80-20 cross-validation scheme was used to split the dataset into training and testing sets, while the training set was further divided into 80-20 for training and validation, respectively. In other words, the ratio of training, internal validation, and testing is 16:4:5.

We aimed to use the simplest convolutional network architecture for our QC model, depicted in Figure 2A.

**Figure 2.**
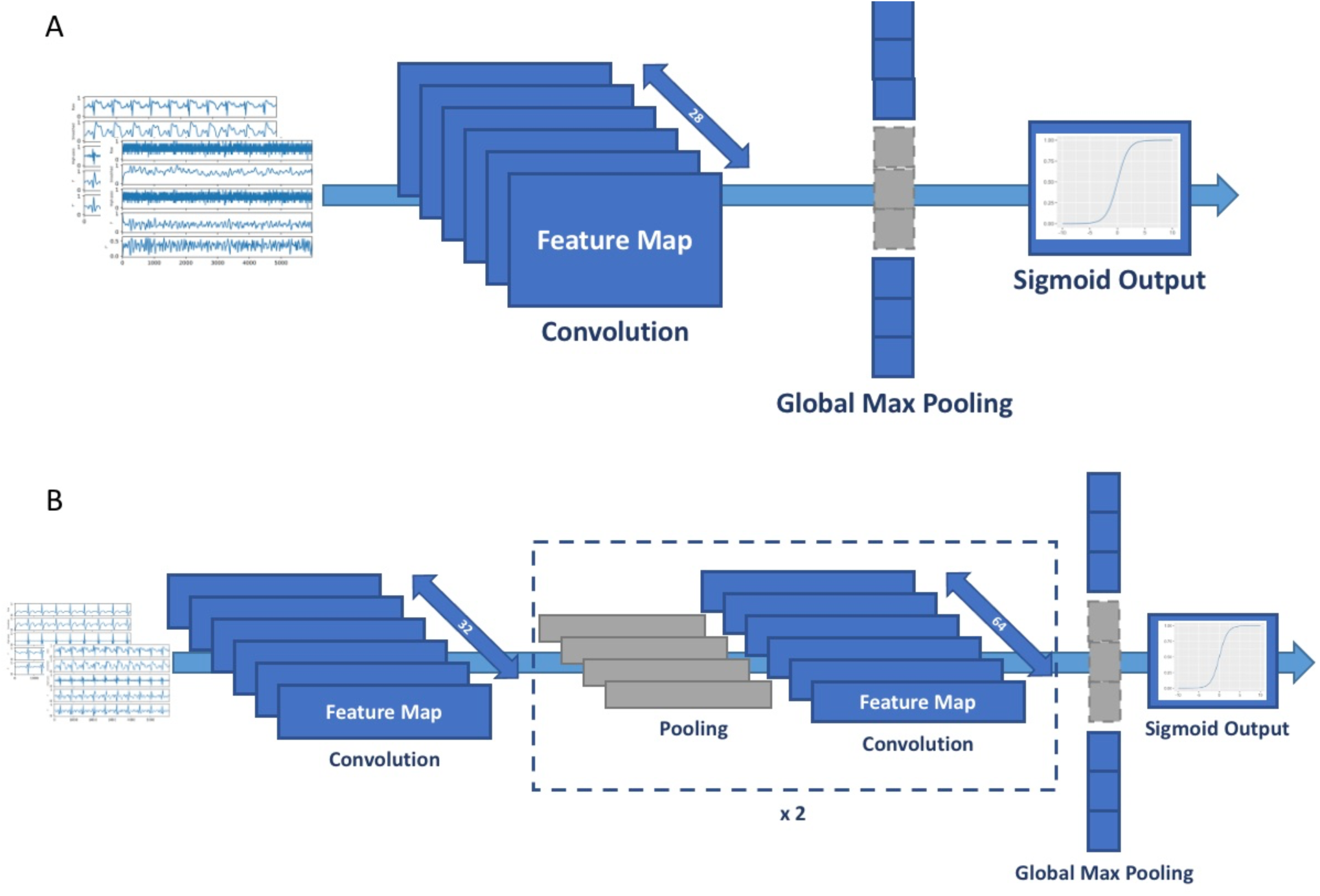
Schematic diagrams of CNN models. A. QC model, and B. stroke model.

The model consisted only of one convolutional layer followed by a fully connected layer (global max pooling). Below are the major model parameters:

- Kernel (filter) size was 20.
- Stride was 10.
- Input layer dimension was 6,000×5.
- The number of filters (feature maps) was 28.
- Rectified linear unit (ReLU) was the activation for the convolutional layer.
- The activation for the fully connected layer was a sigmoid function.
- A dropout layer, with a rate of 50%, was added between the convolutional layer and the fully connected layer to prevent overfitting.
- The loss function was cross-entropy.
- The metric of loss was accuracy.

We used 50 epochs with batch size 30 for model fitting. The model achieved an average of 95% accuracy, where the true positive rate for low- and high-quality segments were 91% and 98%, respectively.

### Stroke Model

The stroke model is similar to the QC model as they are both based on a one-dimensional convolutional neural network model. However, due to the complexity of ECG patterns, the stroke model required additional layers to represent diverse and complex patterns, meaning more training parameters (Figure 2B). Below are the major model parameters:

- Kernel (filter) size was 50.
- Stride was 8.
- Rectified linear unit (ReLU) was the activation for the convolutional layer.
- Input layer dimension was 6,000×5.
- In the first CNN layer, the number of filters was 28.
- In the second and third CNN layers, the number of filters was 64.
- A global maximum pooling layer was used to flatten the last CNN layer into a 64 unit long one-dimensional tensor.
- The activation for the fully connected layer was a sigmoid function.
- A dropout layer, with a rate of 50%, was added between the convolutional layer and the fully connected layer.
- The loss function was cross-entropy.
- The metric of loss was accuracy.

We used 50 epochs with batch size 20 for model fitting. A sample of 12,000 subjects (5,000+ stroke and 6,000+ control) was randomly selected to train the stroke model. Similar to the QC model, the dataset was randomly split into training, internal validation, and testing in the ratio 16:4:5. Note that sample size does influence model accuracy, and its effect will be discussed in the Results section.

To assess the model accuracy, we called the model to make predictions of a randomly selected sample of 1,000 subjects, repeated 20 times. The stroke model has achieved an average accuracy for control (stroke-free), stroke, and combined 85%, 96%, and 90%, respectively. The average runtime was about 85 ms per sample in a 24-core Intel(R) Xeon(R) CPU E5-2620 0 @ 2.00GHz server.

### Class Activation Map

We used the Gradient-weighted Class Activation Map (GRAD-CAM) algorithm[23] to visualize the weights of our two CNN models. Briefly,

- Recombined the input layer and the output of the final layer of a CNN model into a new gradient model. For our models, the input layer was a 6,000×5 tensor, where 6,000 was the length of the signals and 5 was the number of features. The output of the final layer was a 37×64 tensor, where 37 was the length of the convoluted segment, and 64 was the number of filters.
- Invoked the gradient model to predict the label of an input.
- Calculated the gradients of the weights of the gradient model registered during the prediction. The gradients resulted in a 37×64 two-dimensional tensor, the same as the output layer above.
- Calculated the average gradient per filter, giving rise to a one-dimensional tensor of length 37.
- Rescaled/stretched the 37 average gradients into a tensor with the same dimension as the input tensor, i.e., a 6,000 long one-dimensional tensor.
- Generated a heatmap by normalizing the gradient values in the range zero and one.

Two examples of CAMs based on the QC model can be found in Figures 3C and D. The darker the color, the higher is the gradient value or weight.

**Figure 3.**
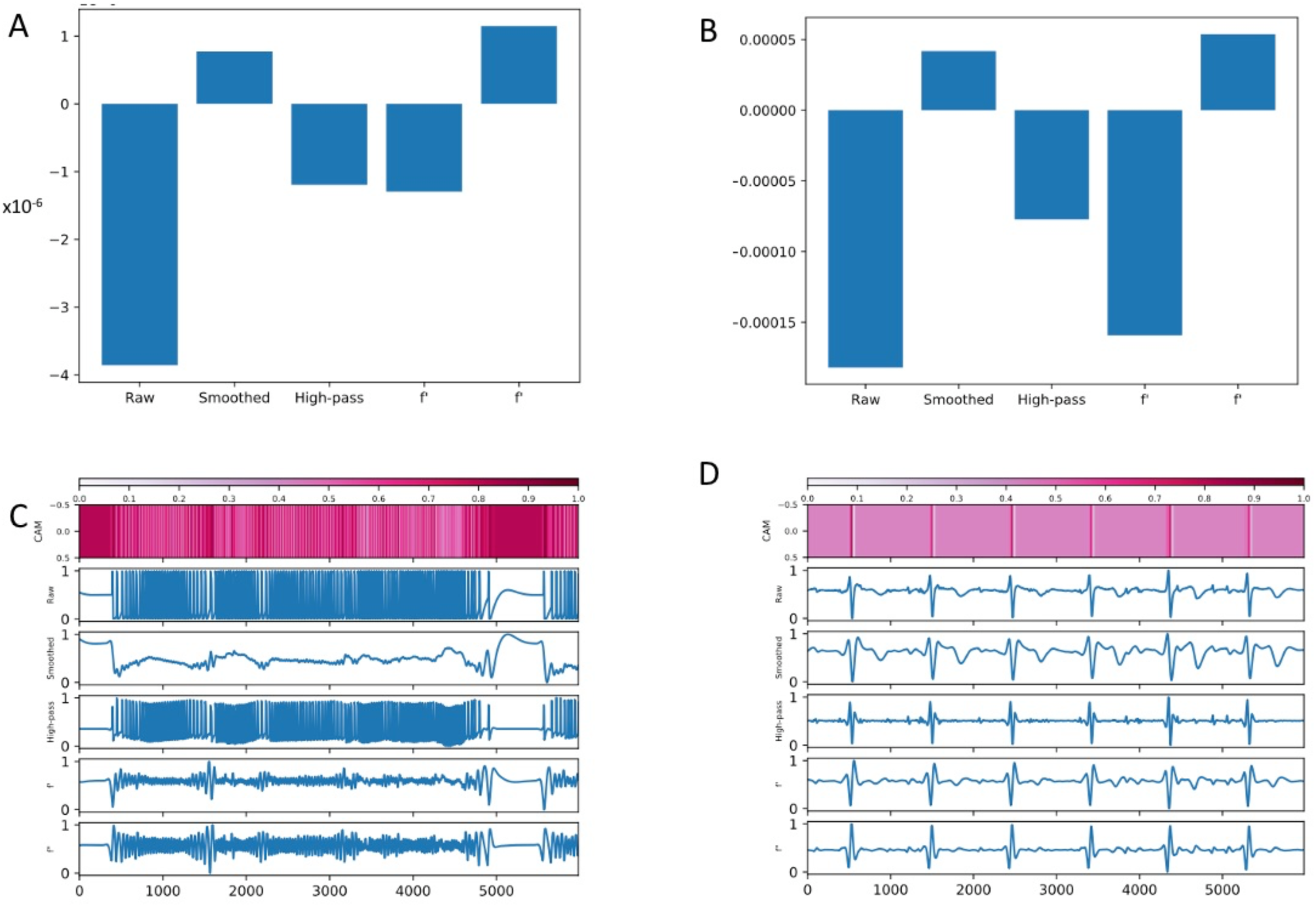
Class Activation Map by GRAD-CAM algorithm. A-B show two examples of gradient importance. A is obtained from a bad-quality segment and B is from a high-quality segment. C-D. CAM heatmaps generated by the GRAD-CAM algorithm. The darker the color, the higher is the weight.

### Gradient Importance

It is helpful to understand the contribution of different features evaluated by the model in making a prediction. We used the following steps to calculate the importance of each feature, which is similar to the GRAD-CAM method:

- Invoked the QC or stroke model to predict the label of an input.
- Calculated the gradients of the weights of the model registered during the prediction. The gradients resulted in a 6,000×5 two-dimensional tensor, the same as the input layer of the model.
- Calculated the average gradient per feature, resulting in five values, one for each feature.
- Plotted the five gradients in a bar chart as shown in Figures 3A and B. The larger the value, the more important is the feature in contributing to the prediction.

### High-Scoring Segments Characterization

To understand patterns learned by the stroke model in distinguishing stroke from stroke-free ECGs, we identified fragments of an input ECG segment where their weights (gradient values) assigned by CAM were higher than 0.8. Recall that weights are in the range of zero and one. Such fragments are called the high-scoring segments (HSSs). We note however that the minimum weight threshold 0.8 was arbitrarily defined as it has produced relevant findings.

Additionally, the position of these HSSs in the PQRST complex could reveal important information about the effect of stroke on ECG. Thus, we characterized each HSS in terms of its ECG waveform. For background, a heartbeat is displayed as a PQRST complex in an ECG. A PQRST complex is comprised of six basic waveforms in this order: P (P-wave), PR (PR segment), QRS (QRS complex), ST (ST segment), T (T-wave), and U wave. As the tool we used to detect these waveforms does not handle U wave, U wave was left out in the analysis, leaving only five basic waveforms. Besides, we created three additional waveforms: W, M, and U. A W waveform indicates an HSS spans a complete or a partial PQRST complex. For example, (P, PR), or (QRS, ST, T). The M waveform denotes an HSS comprised of multiple waveforms but not follows the P, PR, QRS, ST, and T order. For example, M waveform can be (ST, T, P) or (PR, T). Lastly, U stands for unknown.

Neurokit2[24] was used to identify the five basic waveforms. For our dataset, the tool had never detected Q and S waves. Thus, this analysis included only P, QRS, and T waves.

### Pairwise HSS Analysis

As discussed above, HSSs with their associated waveforms were identified in each training or testing sample. We represented HSS waveforms using a single letter as below:

- P for P wave
- R for QRS complex
- T for T wave
- V for PR segment
- W for a complete or a partial PQRST complex
- X for ST segment
- M for multiple waveforms
- U for unknown

For example, seven HSSs have been identified in a sample with these waveforms: M, W, W, W, T, W, and ST. We used this sequence “MWWWTWX” to represent the waveforms.

A sliding window of size two was used to count the occurrences of a pair of neighboring waveforms. For the previous example, these are the six pairs of waveforms: MW, WW, WW, WT, TW, and WX.

We randomly selected 5000 high-quality segments and used the stroke model to predict their classes. For correctly predicted segments, we used the GRAD-CAM model to determine HSSs, followed by waveform identification. As a result, pairs of waveforms were generated and tallied. We repeated this process ten times.

Next, we calculated a z-score for each pair of waveforms by *z* = (*x* − *u*)/*s*, where x is the observed occurrence of a waveform pair, u is the mean occurrence, and s is the sample standard deviation of occurrence. u and s were calculated by bootstrapping where a waveform sequence was shuffled 1000 times. At each time, count the occurrence of waveform pairs in the shuffled sequence. And then, we ranked waveform pairs by descending z-scores. In addition, rank and z-score disparities were calculated for each waveform pair as defined below:

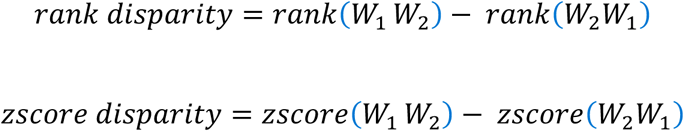

*W*_*1*_ and *W*_*2*_ are the single-letter representation of waveforms.

### Extraction of Individual Waveform

We built six additional stroke models using individual waveform (instead of the entire 6000-ms segment), and the combination of them, i.e., P, QRS, T, PR-interval, QT-interval, and PQRST complex. As above, Neurokit2 was used to identify the locations of different waveforms in high-quality samples. Once the locations were known, a custom Python program was developed to slice them, forming the training and testing datasets for model construction. The same stroke 1D CNN model (Figure 2B) and meta-parameters were used, except the stride parameter was reduced to 2 as segments of individual waveform were much shorter (250 or 750 ms) than 6,000 ms used in the original stroke model.

### Statistics and Machine Learning Software

Statistical analyses were performed using R. The two CNN models described here were developed using Python 3, Tensorflow (version 2.3.0), and Keras (version 2.4.3). We also developed peripheral programs for WFDB to raw signals conversion, random segments sampling, segment visualization, and ECG feature extraction. These programs are available upon request.

## Results

### Overall Workflow

Figure 4 presents the workflow of our stroke model development.

**Figure 4.**
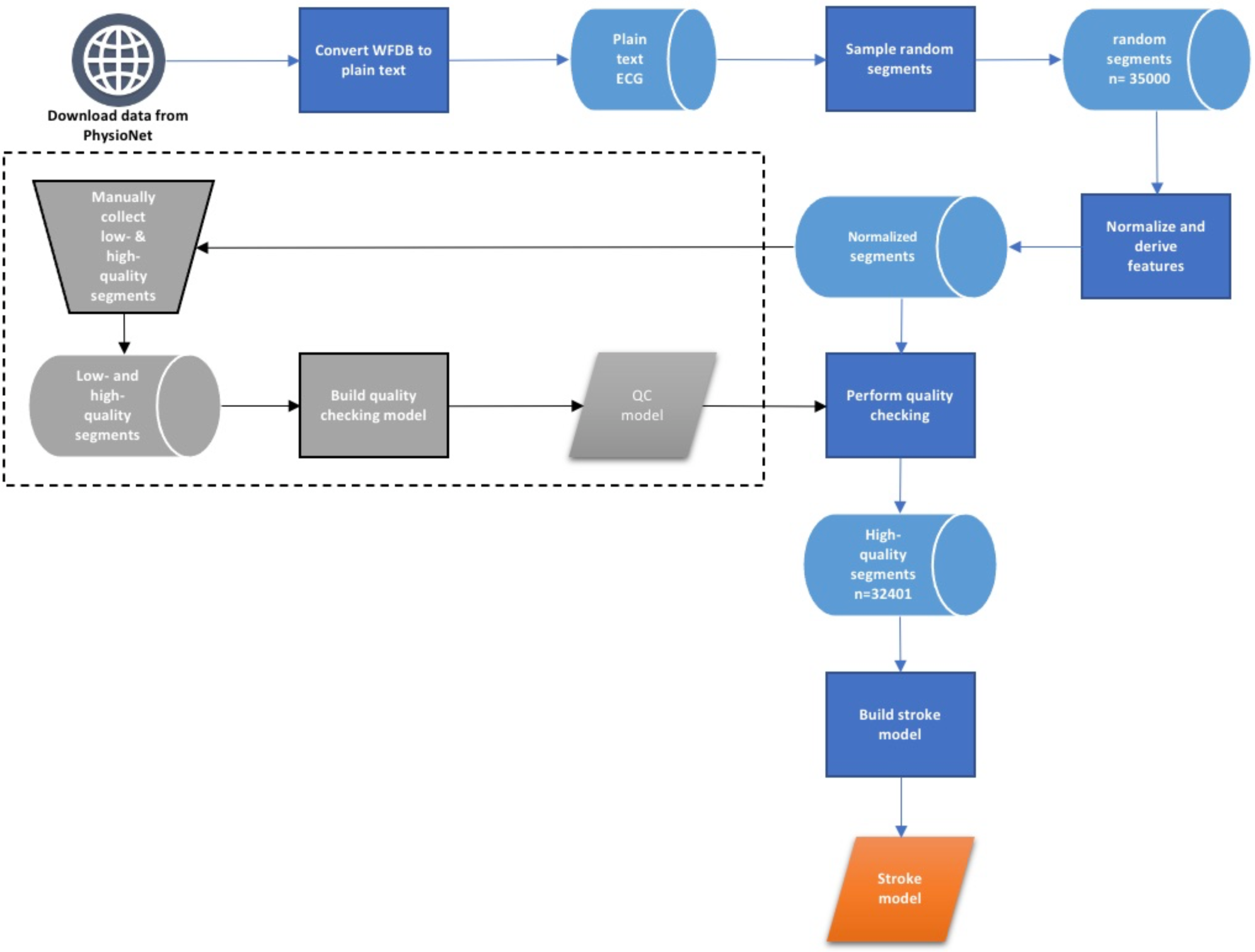
Workflow of the stroke ECG model building process.

Below are the major steps involved in such a model:

- Download the ECG dataset shared by the Cerebral Vasoregulation in Elderly with Stroke study hosted in PhysioNet.
- Convert ECG data from WFDB format to plain text.
- Randomly sample 35,000 ECG segments, each with a length of 6,000 milliseconds, for training, internal validation, and testing purposes.
- Normalize sampled ECG segments and derive four additional features.
- Build a quality checking model for removing low-quality ECG segments before using them for training the stroke model.
- Construct a stroke model to differentiate ECGs originated from post-stroke and stroke-free subjects.

Additional details for these steps are discussed below.

### Quality Checking

While the quality of ECG signals in the downloaded dataset is generally good, it is not immune from sporadic anomalies, as illustrated in Figure 1B. We are set to clean the dataset by a one-dimensional convolutional neural network (1D CNN) to weed out low-quality samples before using it to train the stroke model. We call it the quality checking (QC) model.

Based upon 900+ manually labeled low- and high-quality segments, the QC model has achieved overall 95% accuracy on average, where the correct prediction rate of low-quality and high-quality segments are 91% and 98%, respectively. As model overfitting hampers generalization, we monitored both training and validation losses. Figure 1D depicts a steadily declining plot, suggesting that no concerning overfitting issues exist.

Besides accuracy, interpretation is equally important in assessing the utility of the model. Next, we set to ask what patterns were captured by the model to distinguish low-quality segments from high-quality segments. We used the Gradient-weighted Class Activation Map (GRAD-CAM) method to visualize data points weighted heavily by the model. Two aspects were considered: gradient importance and CAM heatmap. Figures 3A-B are examples of the gradient importance of features for the input signals.

The higher the value, the more important is the feature. For these two examples, smoothed signal and the second derivative contribute positively to the model for that instance. It is noteworthy that what features are considered important by the model vary from sample to sample, unlike other machine learning methods such as logistic regression, in which the model coefficients are constant with respect to input sample.

The class activation map (CAM) complements gradient importance in visualizing indicative regions that the model focused. The top subpanel of Figures 3C-D show two CAMs, one for a low-quality segment and the other for a high-quality segment. The darker the color, the higher is the weight. Noticeably, the QC model assigns high weights to a broad region in a low-quality segment. On the contrary, the model hones in on the repeated occurrences of QRS complexes of a high-quality segment. In other words, the QC model recognizes that a cyclic spike is the signature of a high-quality segment. Intriguingly, the model has learned to differentiate different cyclic frequencies, i.e., remarkably high-frequency patterns pertaining to low-quality signals (the middle section of Figure 3C) from physiological-sensible cyclic patterns illustrated by normal heartbeats.

### Stroke Model Assessment

We used the QC model to filter the training dataset before using them for building the stroke model. Out of the 34,200 randomly selected, non-overlapping ECG segments, 1,799 (about 5%) were classified as low-quality, yielding 32,401 segments for stroke model building. We assessed the stroke model in four perspectives: i) accuracy, ii) low-quality samples prefiltering, iii) segment length, and iv) sample size.

i. Model accuracy. The stroke model was used to make predictions for 1000 randomly selected subjects, repeated 20 times. The median accuracy for control (stroke-free), stroke, and overall are 85%, 96%, and 90%, respectively (Figure 5A). Alongside, the area under the receiver operating characteristic curve (AUROC) was used to measure accuracy where 0.5 is random and 1.0 is perfect. Our model has achieved 0.95 (Figure 5B). We also monitored overfitting by examining the disparity between training and validation loss and accuracy (Figure 5C). Because both validation curves follow the downward trend of the corresponding training curves, overfitting is under control.
ii. Low-quality samples pre-filtering. In the above, we argued that removing low-quality data can improve the stroke model’s accuracy. Using a semi-automatic approach, we developed a quality checking model for removing low-quality segments before training the stroke model. To justify that such a step is necessary, we planted 5% to 15% of low-quality segments into the training samples while constructing the stroke model. As shown in Figure 5D, the model’s accuracy declines according to the increasing percentage of low-quality segments, substantiating the notion of low-quality data prefiltering.
iii. Segment length. Do longer segments contribute to higher accuracy? We trained and tested the stroke model with segment lengths 2,000 ms, 4,000 ms, 6,000 ms, and 10,000 ms (Figure 5E). Their mean accuracies were compared by one-way ANOVA, followed by post-hoc Tukey’s HSD test. The only significant difference detected was 2,000 ms versus 4,000 ms (p=2.133×10^-4^, at 95% C.I.), with the accuracy of 4,000 ms was more superior to 2,000 ms. Giving the median accuracy of 10,000 ms is low than 6,000 ms (not statistically significant), the remaining choices are 4,000 ms and 6,000 ms. As a longer segment may ease interpretation as illustrated by CAM, 6,000 ms should suffice for this study.
iv. Sample size. ECGs carry complex cardiac signals that entail essential physiological information. Therefore, we investigated the adequate number of training samples to attain the best possible accuracy. The sample sizes tried are 6,000, 10,000, 12,000, and 14,000. Accuracy rises from 80% to almost 90% according to increasing sample sizes (Figure 5F). One-way ANOVA test followed by post-hoc Tukey’s HSD test suggested no difference among sample sizes 10,000, 12,000, and 14,000. Therefore, a sample size of 14000 was used for this project.

**Figure 5.**
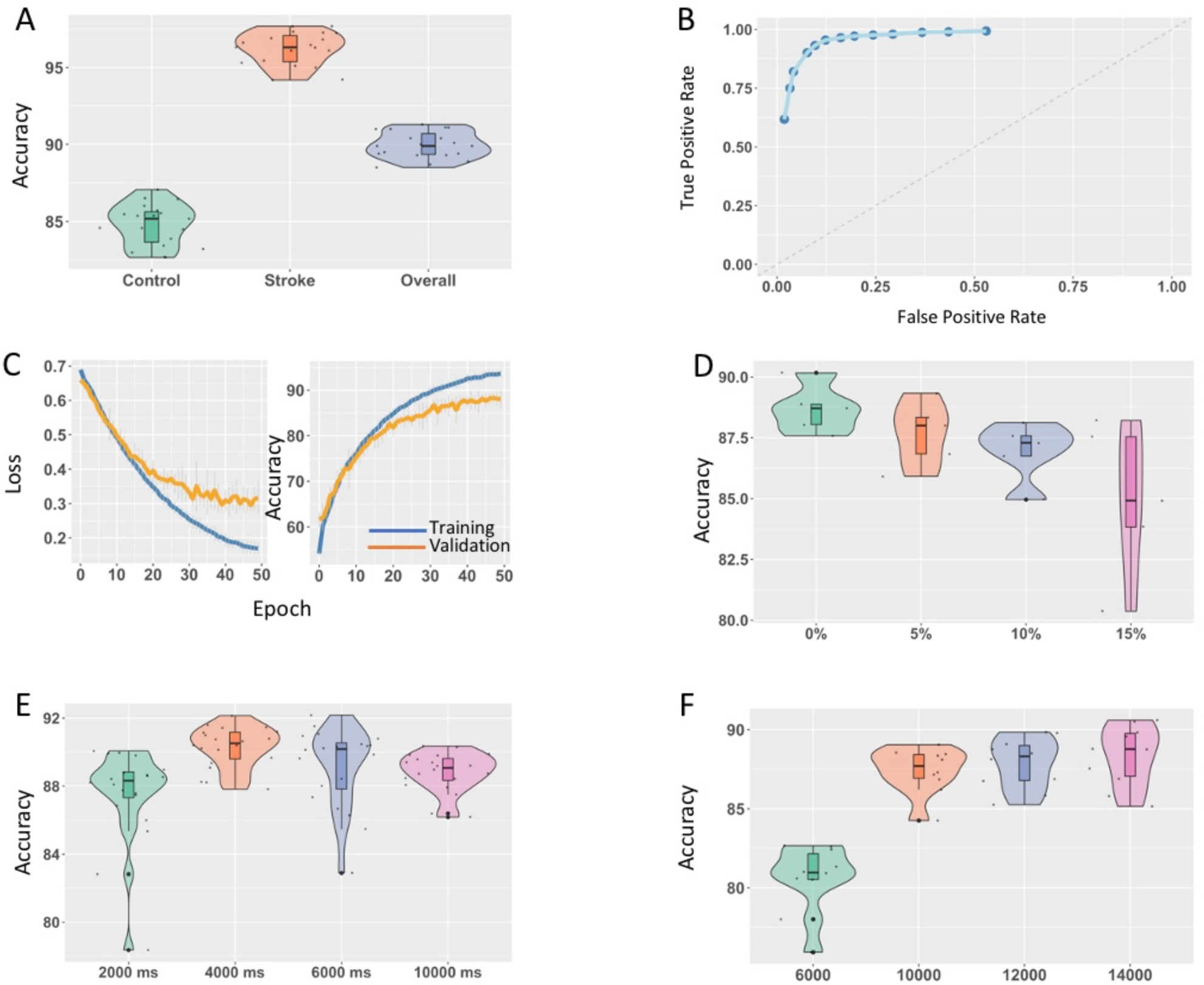
Stroke model assessment. A. model prediction accuracy results from 20 repeated tests. The model made predictions for 1,000 subjects with control (stroke-free) to stroke almost 1:1. B. Receiver Operating Characteristic (ROC) curve of the stroke model based on the test results from A. C. Loss and accuracy curves for training and validation. D. Evaluating the effect of low-quality data on accuracy. E. Comparing accuracy by segment length. F. Assessing the effect of sample size on model accuracy.

### Interpretation of Stroke Model

While CNN and deep neural networks alike are superior in prediction accuracy, the model is opaque in convincing users features or patterns harnessed by the model in make a prediction or classification. To elucidate features learned by our stroke model, we extracted high-scoring segments (HSSs) guided by the weights computed by CAMs. HSS is a segment in the sample where the CNN model weights highly in differentiating the two classes. These highly weighted segments are illustrated in deep red in Figure 6A.

**Figure 6.**
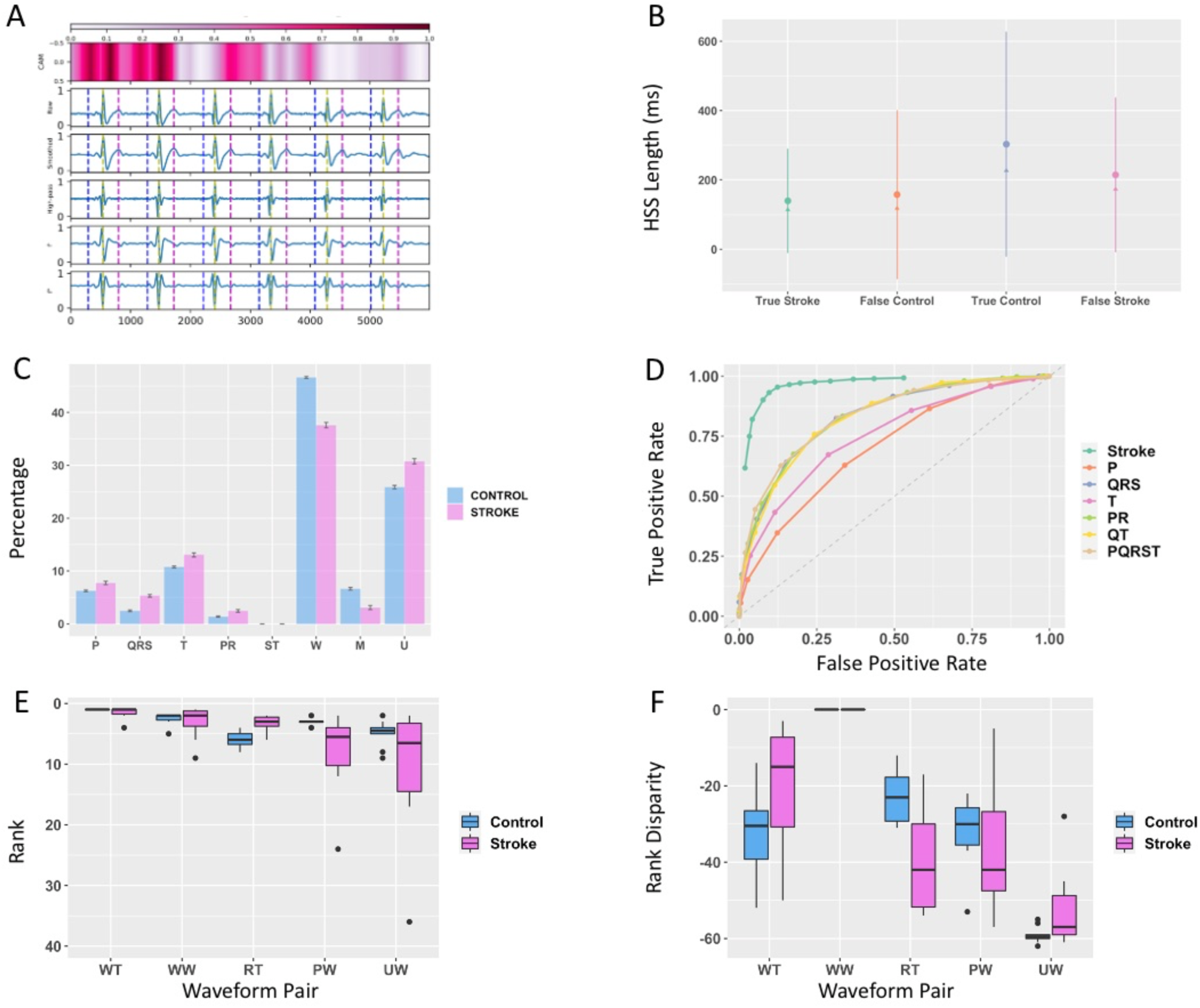
Model interpretation. A. CAM heatmap a high-quality ECG segment. The horizontal heatmap at the top was produced by GRAD-CAM algorithm. It represents the weights assigned by the stroke model for the signals. The darker the color, the higher is the weight, meaning that it contributes more in determining the label. The vertical dash lines mark the P-peak (blue), R-peak (yellow), and T-peak (magenta). B. It shows the range (mean± s.d.) of high-scoring segments (HSSs) by prediction outcome. Mean and median are denoted by a dot and a triangle, respectively. C. The distribution of HSSs by basic waveforms: P (P-wave), QRS (QRS complex), T (T-wave), PR (PR segment), ST (ST segment), W (the whole or partial PQRST wave), M (multiple types), and U (unknown). D. ROC curves of models built by individual waveform or a combination of basic waveforms. The top left ROC belongs to the full stroke model, same as the ROC in Figure 5B. AUROC of P, QRS, T, PR interval, QT interval, and PQRST are 0.62, 0.78, 0.68, 0.79, 0.78, and 0.79, respectively. E. The top five ranked waveform pairs by z-score. Rank 1 means the largest z-score. F. Rank disparity of the waveform pairs from E. Rank disparity is defined as the rank difference between W_1_W_2_ and W_2_W_1_ where W_1_ and W_2_ are the first and the second waveform of a waveform pair.

We compared the lengths of HSSs among four prediction outcomes, i.e., true stroke, false control, true control, and false stroke, as shown in Figure 6B. Note that stroke-free is labeled as control. All pairs show significant differences (at 95% C.I.) in length except for true stroke vs. false control according to the pairwise t-test.

Besides length, we are keen to associate the PQRST waveforms with these HSSs and their distribution (Figure 6C). Apart from PR and ST segments, the prevalence of each waveform is statistically different (at 95% C.I.) between the post-stroke and the stroke-free. It suggests that not all waveforms are contributed equally in class label prediction.

With regard to correctly predicted samples, the model paid attention to small fragments (mean 140 ms, median 114 ms) for the post-stroke instead of longer fragments (mean 303 ms, median 227) in the stroke-free (Figure 6B). But they are still within a heartbeat for both classes. This observation concurs with the PQRST waveform distribution in Figure 6C as the most frequent type is W, i.e., the whole or partial PQRST complex, where its length is approximately 400 ms long. As M waveform occupies <5% of all kinds, the model rarely focused on inter-heartbeat patterns. This result is consistent with the HSS length analysis since the interval between two adjacent heartbeats (∼900 ms) is much longer than 300 ms observed. Notably, the unknown (U) occupies 30% of the HSSs. High signal-to-noise is ostensibly distorted the PQRST complex, especially for P and T waveforms, posting a challenge to waveform detection. Moreover, when the whole or a partial PQRST was not used (i.e., not W), the next preferences for the model are individual waveforms (P, QRS, or T). Such a switch appeared in both classes, but it is more pronounced in the post-stroke. Additional evidence is revealed in the section “HSS Pairs Dependency” below.

For post-stroke cases misclassified as stroke-free (false control), the model mistakenly focused on short fragments (about 158 ms, slightly longer than the true stroke), leading to erroneous decisions. As discussed above, a longer fragment is needed in discerning a stroke-free segment; missing integrated information of the entire PQRST complex is probably the source of errors. With this in mind, it could allude to ECG segments not exhibiting pathogenic ECG patterns as a post-stroke subject’s ECG may display anomalous signals all the time.

Regarding stroke-free segments mistakenly classified as post-stroke (false stroke), the model did examine longer fragments (215 ms) than true stroke (140 ms) cases. Still, it is only ∼70% of the average HSS length from the true control (303 ms). The error is likely due to the skipping of vital signals presented in a broader region in deciding the class label.

### Stroke Models Based on Individual Waveform

Based on the analysis above, individual waveforms P, QRS, and T seem to contain informative signals that complement the W waveform. To assess the extent of contribution alluded to individual waveform, we built six additional stroke models based upon individual waveform and their combinations, including P, QRS, T, PR interval, QT interval, and the whole PQRST complex. To compare their accuracy with the original stroke model, we plotted their ROC curves as shown in Figure 6D. Unsurprisingly, the full-blown stroke model performs the best, with at least 17 percentage points lead (AUROC 0.95 from the stroke model versus 0.78 from the QRS-model, see also Figure 6C legend) compared with other models.

Intriguingly, the QRS-model had outperformed the T- and P-model, although more HSSs are associated with the P and T waveforms (6-8% and 11-13%, respectively) than the QRS waveform (3-5%), as shown in Figure 6C. Overall, the P-model is the worse. And the QRS-model is as competitive as the PR, QT, and PQRST models. In short, this result has discovered the differential informativeness of individual waveform in classifying post-stroke from stroke-free ECGs. More importantly, it is perplexing to us why the more frequently used waveforms do not contribute to better model accuracy. Such an observation leads us to explore if the model synergically combines adjacent HSSs to improve prediction accuracy.

### HSS Pairs Dependency

We asked the question if waveforms pertaining to adjacent HSSs are independent or not. We took a bootstrapping approach to quantify the co-occurrence of waveforms W_1_ and W_2_ in a pair (W_1_W_2_) versus random. In short, 5,000 high-quality segments were sampled, followed by class label prediction. HSSs and corresponding waveforms were identified for correctly predicted samples. The occurrence of waveform pairs was tallied. Finally, z-scores for waveform pairs were computed and ranked. The higher the z-score, the more probable W_1_ and W_2_ are coupled rather than independently assorted. Besides, the rank and z-score disparities were computed for each waveform pair (W_1_W_2_). The Materials and Methods section provides a detailed description of the procedure.

As shown in Figure 6E, waveform pair WT (W followed by a T wave) is consistently ranked the first in ten repeated trials in both control and stroke. Additionally, its rank disparity is negative (-30 in control and -15 in stroke), implying the existent of intrinsic order in which W followed by T is preferred (ranked high) by the stroke model instead of the reverse. All top five waveform pairs exhibit such asymmetric coupling except for WW. It is found that WT, WW, RT, PW, and UW occur in 19%, 37%, 2%, 10%, and 34%, respectively, of all samples. These findings have answered the question why more frequent waveforms, such as P and T, alone do not improve model accuracy as they were combined with W synergistically. With this in mind, the W of WT and PW may represent the beginning and ending of a PQRST complex, respectively.

If this argument is valid, why PQRST-model does not perform as good as the full-blown stroke model? We speculate that there are informative signals before and after the core PQRST complex and these signals were not characterized by Neurokit2, such as the U-wave. This point is ascertained by the fact that UW is the fifth-highest waveform pair (Figure 6 E-F) and U type makes up 25-30% of all waveform types (Figure 6C). In summary, there are weak unknown but illuminating signals characterize either the post-stroke or the stroke-free. Further improvement of waveform detection tools is needed to confirm our findings.

## Conclusions

We have presented an effective model that can distinguish ECGs from post-stroke and stroke-free with ∼90% accuracy and AUROC 0.95. Our study is distinct from previous studies as we have analyzed the underlying patterns captured by the convolutional neural network model. By using the GRAD-CAM algorithm, we were able to unravel waveform patterns ascribed to the model’s accuracy. Results suggest that solely the core PQRST complex is inadequate to characterize a post-stroke ECG. Instead, the model has taken into signals precede or after the core PQRST complexes in class prediction. Such an understanding is essential in alluding to cardiac issues experienced by the post-stroke, supporting clinicians in making therapeutic decisions. It is noteworthy that long QT is associated with inherited cardiac arrhythmia, a risk factor of ischemic stroke, however we found no statistical difference between post-stroke and stroke-free subjects (data not shown).

That said, our work has several limitations. Data paucity is a challenge as the dataset we acquired contains only a small number of elderly subjects (n=91). The etiology of stroke is complex. Thus, we expect to see diverse ECG patterns from the post-stroke. Although the overfitting of our model is under control (Figure 5C), expanded recruitment of subjects can definitely safeguard model generalization. As atrial fibrillation is a known risk factor of cardioembolic stroke, ECG analysis presented in this study may play a more relevant role in this subtype than other stroke subtypes such as small vessel disease. Thus, stratifying stroke subtypes in the future study can improve prediction by pinpointing subtype-specific ECG patterns.

Additionally, there are two issues presented in the public ECG dataset. First, the ECG segments were randomly selected where the corresponding activities performed by subjects were unknown, i.e., whether or not they were recorded during resting or walking, etc. The missing of such information could introduce uncertainty for the model. Second, since the study required uninterrupted measurements over a 24-hour period, ECGs were recorded by only two leads of the portable device. Admittedly, if 12-lead ECGs were available, our model is a capable of learning the nuance of ECGs between the post-stroke and the stroke-free. However, this is the only public available stroke ECG dataset known to the authors.

Moreover, U-wave was not taken into account as it is obscure to the detection software. Although no study has alluded stroke to U-wave at present, our pairwise HSS analysis suggests that subtle but unknown signals precede or after the core PQRST complex may be distinctive to the post-stroke. Thus, a more sensitive waveform detection method is needed to fit such a gap.

Lastly, this is a retrospective study. The clinical value of the current model is limited. If longitudinal data is available in the future, our model can be enhanced to predict the risk of stroke, especially for young people (<65 years) with a lifestyle prone to stroke.

Given stroke’s high disability-adjusted life year, the 1D-CNN model incorporated with GRAD-CAM is a promising deep neural network framework that can be applied in clinical environment, mitigating disease burden.

## Data Availability

Dataset was obtained from another research group: https://physionet.org/content/cves/1.0.0/

## Acknowledgements

We thank Prof. Bradley C. Antanaitis and Dr. Yiu-man Wong from the Physics Department of Lafayette College for the insightful discussion of ECG analysis.

